# Too much information? Asian Americans’ preferences for incidental brain MRI findings

**DOI:** 10.1101/2023.04.17.23288629

**Authors:** Karthik Kota, Alice Dawson, Julia Papas, Victor Sotelo, Guibin Su, Mei-Ling Li, Woowon Lee, Jaunis Estervil, Melissa Marquez, Shromona Sarkar, Lisa Lanza Lopez, William T. Hu

## Abstract

**INTRODUCTION:** South Asian (SA) and East Asian (EA) older adults represent the fastest growing group of Americans at risk for dementia, but their participation in aging and dementia research has been limited. While recruiting healthy SA older adults into a brain health study, we encountered unexpected hesitancy towards structural brain MRI analysis along with some stigmatizing attitudes related to internal locus of control (LoC) for future dementia risks. We hypothesized that support for MRI-related research was influenced by these attitudes as well as one’s own MRI experience, perceived MRI safety, and concerns for one’s own risks for future dementia/stroke.

**METHODS:** We developed a brief cross-sectional survey to assess older adults’ MRI experiences and perceptions, desire to learn of six incidental findings of increasing health implications, and attitudes related to dementia as well as research participation. We recruited a convenience sample of 256 respondents (74% reporting as 50+) from the New Jersey/New York City area to complete the survey, and modeled the proportional odds (P.O.) for pro-research attitudes.

**RESULTS:** 77 SA and 84 EA respondents were analyzed with 95 non-Asian adults. White (P.O.=2.54, p=0.013) and EA (P.O.=2.14, p=0.019) respondents were both more likely than SA respondents to endorse healthy volunteers’ participation in research, and the difference between White and SA respondents was mediated by the latter’s greater internal LoC for dementia risks. EA respondents had more worries for future dementia/stroke than SA respondents (p=0.006), but still shared SA respondents’ low desire to learn of incidental MRI findings.

**DISCUSSION:** SA and EA older adults had different attitudes towards future dementia/stroke risks, but shared a low desire to learn of incidental MRI findings. A culturally-appropriate protocol to disclose incidental MRI findings may improve SA and EA participation in brain health research.

**Color printing:** Please have figure one and two be in color; figure three is in black and white

## 1. Introduction

Asian Americans represent the fastest growing racial group in the U.S., nearly doubling in number from 2000 to 2019. Asian Americans report ancestry from East Asia, the Indian Subcontinent (South Asia), and South East Asia, with over half of older Asian Americans living in California, New York, Texas, New Jersey, and Washington.[1] Health record studies suggest older Asian Americans to have lower prevalence of dementia diagnosis than older White adults,[2] but whether this is due to a difference in dementia risk, detection, or stigma remains controversial.[3] This also contradicts the observation that standardized dementia prevalence is comparable in Asia (5.63% in South to 7.15% in Southeast Asia), Europe (4.65% in Central to 6.67% in Western Europe), and North America (6.77%),[4] but Asian immigrants to the U.S. can differ from their counterparts in Asia according to educational ascertainment,[5] socioeconomic status,[5] and health behaviors.[6] The most common causes of dementia in older Asian Americans are also poorly understood. Among the Asian immigrant groups, South Asian (SA) adults – primarily from India but also Bangladesh, Bhutan, the Maldives, Nepal, Pakistan, and Sri Lanka – are noted in North American studies to have disproportionate atherosclerotic and cardiovascular disease risks.[7] This potentially suggests greater vascular than degenerative contribution to dementia among older SA adults; however, this assumption is complicated by differential genetic risks, acculturation,[8] and greater stroke mortality among all Asian subgroups than White adults.[9] Due to potential family objection to post-mortem neuropathological examination related to religious reasons or knowledge,[10] ante-mortem biomarker studies are most likely to shed insight into etiologies and mechanisms underlying dementia in older SA adults.

Structural brain MRI is a common clinical and research tool to detect cerebrovascular disease and neurodegeneration. Most discussions have focused on its safety in, rather than acceptability to, participants.[11] This is especially relevant in aging studies, where older adults face serial high field or one-time ultra-high field MRI analysis of increasing duration and complexity for research.[12] One recent study showed aggregated Asian American respondents as likely as other groups to express willingness to undergo research brain MRI,[13] but less is known about disaggregated East Asian (EA) and SA adults’ willingness. Limited literature shows low knowledge of the non-radiation nature of MRI in China[14] and nocebo effects of overly clinical MRI reports on patient outcomes in India.[15] How these factors translate to North America remains largely unknown.

Beyond willingness to participate, incidental findings on brain MRI are identified in 1.7-4.3% of research scans.[16] Disclosure and follow-up of incidental research MRI findings continue to be a relatively under-examined topic,[17-19] with mainstream protocols in the U.S. and U.K. following regulatory guidelines consistent with Western legal and ethical framework.[19-21] At the same time, their cultural relevance may be overlooked when diverse older adults are recruited into aging research. While these cultural factors may get less attention in clinical settings, their impact on research participation and trust in researchers – especially among diverse older adults – remains unexplored.

In the course of recruiting diverse older adults into a prospective memory and aging study in the New Jersey/New York City (NJ/NYC) area, we encountered significant concerns from potential SA participants about the study’s MRI component. Although MRI was previously considered a broadly acceptable non-invasive modality in the U.S. and U.K., prior studies have not included large numbers of older SA adults.[22] To better understand factors associated with MRI avoidance in SA older adults, we developed a short survey to examine past MRI experiences (brain or body); desire to learn of incidental findings on brain MRI; and attitudes towards brain MRI safety, dementia, and research participation.

For attitude-related questions, we emphasized several shared beliefs among SA older adults related to focused on fear of future dementia since it is a known risk factor affecting dementia research participation.[23, 24] Locus of control (LoC) refers to the belief that an individual’s actions affects their health outcome, and those with a high internal LoC tend to utilize less health care.[25] Relevant to this study, SA adults have been found to participate less in health activities due to self-perceived high LoC.[26] We hypothesized that SA older adults have low prior exposure to MRI, a low wish to learn of incidental MRI findings, the belief that MRI is harmful, and high internal LoC related to brain health, which lead to their low willingness to participate in MRI-related research. Since shared as well as distinct experiences (e.g., immigration) and cultural beliefs among different U.S. Asian ethnic groups shape perception and stigma associated with dementia and brain health,[27] we additionally recruited EA and non-Asian older adults to test this hypothesis.

## 2. Materials and Methods

### 2.1 Consent statement

The anonymous MRI survey did not collect protected health information, and the study was considered exempt human subject research by Advarra Institutional Review Board.

### 2.2 Survey

Our cross-sectional MRI survey collected information on racial/ethnic (option to select from Hispanic, White, Black, East Asian [e.g., Chinese, Korean], South Asian, Other with write-in space) and age categories (<50, 50-59, 60-69, 70-79, 80+); experience with any or brain MRI (yes, no, not sure); perceived prevalence of incidental findings on brain MRI among healthy people (<5%, 5-10%, 15-25%, >50%); and ten Likert-scale questions on attitudes related to research and brain health, administered via paper (n = 156) and online (n = 100). The two research-related questions were: *“Healthy people do not need to participate in medical research, because there is no direct benefit to them”; “After my name is removed from my brain scan, I am comfortable with the idea of scientists [who are not all physicians] I have never met examine the images*.*”* Brain health-related questions included three on dementia risk factors previously mentioned during community meetings with SA older adults *(“A diet low in meat is effective in preventing Alzheimer’s disease and dementia”; “Brain diseases like Alzheimer’s and dementia only happen to people who don’t regularly exercise their brains [e*.*g*., *through reading];” “Only people with diabetes or strokes will develop dementia”)*, two on MRI safety *(“Putting a person in a strong magnetic field cannot cause long-term physical harm”; “Putting a person in a strong magnetic field cannot cause long-term mental harm”)*, and one each on forgetfulness (“Forgetting conversations and appointments I don’t care about is a normal part of aging”), concerns about one’s own future brain health *(“I am afraid that I might have dementia or strokes in the future”)*[28] and dementia-related stigma *(“People with dementia should be separated from society for their and our safety”*) based on their relationships to research participation.[29] Since the meaning of survey-based hypothetical willingness to participate in research has been criticized, we did not include a question on willingness.[30] All statements were examined for wording, clarity, and potential for misinterpretation; piloted in 13 volunteers (seven SA older adults); and revised for clarity (one question). No pilot data were included for subsequent analyses. Surveys were translated into Chinese, Korean, and Spanish through forward and backward translation, followed by proofing by bilingual project scientists fluent in these languages. We did not translate the surveys into SA languages since there were too many to offer equitable representation, and did not have study personnel who were fluent and could guarantee cultural appropriateness and freedom from bias for the surveys. Surveys took three to five minutes to complete.

### 2.3 Participants

A convenience sample of participants was recruited to complete the survey between August 2022 and January 2023. Potential participants were recruited from Rutgers General Internal Medicine Clinic (n=8), Rutgers Neurology Clinic (n=37), community events related to aging and health disparities research (n=64), and direct solicitation through research registries and word-of-mouth referrals (n=147). Participants were eligible if they could read and respond to questions in English, Chinese (simplified or traditional), Korean, and Spanish. We primarily recruited participants who were likely 50 years of age or older. Respondents were told the study’s purpose was to “understand attitudes and knowledge about MRI and dementia.” Participants were not offered incentives.

### 2.4 Statistical analysis

All statistical analysis was conducted in SPSS 28.0 (IBM-SPSS, Armonk, NY). Differences between groups were analyzed by Chi-squared tests for variables of categorical nature (e.g., race/ethnicity, source of recruitment) or regression analysis (e.g., ordinal for belief that healthy people should volunteer, linear for summary measures). There were 28 EA participants responding in simplified or traditional Chinese, and 13 EA participants responding in Korean. EA responding in Chinese or self-identified Chinese ethnicity (n = 50) and EA responding in Korean or self-identified Korean ethnicity (n = 16) were older than EA respondents who responded in English or did not specify ethnicity (n = 18; median age 60-69 vs. <50), but were otherwise similar in MRI experience and attitudes. We thus analyzed all three subgroups together as one EA group. No participant responded to the Spanish survey. 7 respondents reported Other race/ethnicity, and were excluded from analyses according to race/ethnicity.

For attitudes related to research, responses to the two Likert-scale questions on research participation were analyzed as dependent variables in ordinal regression models. Models involving five response categories for healthy people as research participants met the parallel regression assumption. Models involving sharing de-identified MRI did not meet the parallel regression assumption when five categories were used, but did meet the assumption with three categories (disagree, neutral, agree), which may reflect effects of extreme answers. Ordinal outcomes involving these three categories were thus used for sharing de-identified MRI. For logistic and ordinal regression, odds ratio (O.R.) and proportional odds (P.O.) are reported with 95% confidence interval (CI).

For attitudes related to brain health, we performed principal component analysis (PCA) on the eight Likert-scale questions. Principal component (PC) scores were calculated, analyzed across racial/ethnic groups using Analysis of Covariance (ANCOVA), and then analyzed across age categories using ordinal regression analysis. PC scores were also entered into regression models of attitudes on research participation, and PC scores which differed between racial/ethnic groups were additionally entered into mediation analyses.

For attitudes towards incidental MRI findings (Table 2) and regression-based mediation analysis (Table 4), a sample size of 249 (excluding self-identified “Other” respondents) was sufficient to achieve power of 0.91 to detect an effect size of 0.3 at α of 0.05. For comparison of PCs, a sample size of 249 was sufficient to achieve power of 0.88 to detect an effect size of 0.20 at α of 0.05.

## 3. Results

### 3.1 MRI experiences and facts

Among 256 respondents, 33%, 30%, 21%, 7%, and 6% identified as EA, SA, White, Black, and Hispanic respectively. 66 (26%) reported age under 50 (Table 1), and Hispanic respondents were younger than SA respondents (p=0.020). White respondents were more likely recruited via registries and word-of-mouth referral than non-White respondents (100% vs 67%, p<0.001). The respondents were otherwise similar in age and recruitment source.

**Table 1.** Survey respondents’ demographic information, MRI experience, wish to learn of incidental MRI finding, and research-related attitudes. 66 (78%) East Asian participants volunteered ethnicity information or responded in a specific language (Chinese, Korean), and their responses are shown within each ethnic grouping but also as one East Asian group. Median age category in each racial/ethnic group was 60-69 except for East Asian, NOS (<50); Hispanic (50-59); and Other (<50). (* One Hispanic respondent did not provide age)

|  | Total East Asian (n=84) | Chinese or EA responding in Chinese (n=50) | Korean or EA responding in Korean (n=16) | EA, NOS or EA responding in English (n=18) | South Asian (n=77) | White (n=54) | Black (n=19) | Hispanic (n=15) | Other (n=7) |
| --- | --- | --- | --- | --- | --- | --- | --- | --- | --- |
| Age |  |  |  |  |  |  |  |  |  |
| <50 | 22 | 5 | 5 | 12 | 13 | 14 | 5 | 7* | 5 |
| 50-59 | 14 | 9 | 2 | 3 | 7 | 10 | 3 | 0 | 0 |
| 60-69 | 25 | 17 | 6 | 2 | 35 | 16 | 2 | 6 | 1 |
| 70-79 | 18 | 15 | 3 | 0 | 17 | 11 | 4 | 1 | 1 |
| 80+ | 4 | 3 | 0 | 1 | 5 | 3 | 4 | 0 | 0 |
| Had any prior MRI (%) | 42 (50%) | 24 (48%) | 8 (50%) | 10 (56%) | 39 (51%) | 39 (72%) | 13 (68%) | 9 (60%) | 3 (43%) |
| Had a prior brain MRI (%) | 23 (27%) | 13 (26%) | 5 (31%) | 5 (28%) | 16 (21%) | 26 (48%) | 4 (21%) | 5 (33%) | 2 (29%) |
| Wish to learn of incidental MRI findings |  |  |  |  |  |  |  |  |  |
| Entirely benign | 24 (29%) | 17 (34%) | 3 (19%) | 4 (22%) | 27 (35%) | 29 (54%) | 8 (42%) | 12 (80%) | 4 (57%) |
| Past injury, no longer threat | 22 (26%) | 15 (30%) | 2 (13%) | 5 (28%) | 29 (38%) | 31 (57%) | 8 (42%) | 7 (47%) | 4 (57%) |
| Common findings, can improve health | 42 (50%) | 26 (52%) | 8 (50%) | 8 (44%) | 45 (58%) | 33 (61%) | 10 (53%) | 9 (60%) | 5 (71%) |
| Unclear findings, need more testing | 29 (35%) | 15 (30%) | 7 (44%) | 7 (39%) | 33 (43%) | 39 (72%) | 8 (42%) | 10 (67%) | 4 (57%) |
| Serious finding with treatment | 50 (59%) | 25 (50%) | 11 (69%) | 14 (78%) | 44 (57%) | 43 (80%) | 12 (63%) | 12 (80%) | 5 (71%) |
| Serious finding with no treatments | 25 (30%) | 13 (26%) | 4 (25%) | 8 (44%) | 27 (35%) | 35 (65%) | 8 (42%) | 7 (47%) | 4 (57%) |
| Research Q1 | n=78 | n=47 | n=16 | n=15 | n=70 | n=47 | n=14 | n=15 | n=7 |
| Strongly agree | 7 | 5 | 0 | 2 | 11 | 1 | 0 | 2 | 0 |
| Agree | 3 | 1 | 2 | 0 | 6 | 3 | 2 | 1 | 1 |
| Neutral | 11 | 7 | 4 | 0 | 14 | 7 | 4 | 2 | 1 |
| Disagree | 12 | 7 | 3 | 2 | 12 | 10 | 3 | 7 | 1 |
| Strongly disagree | 45 | 27 | 7 | 11 | 27 | 26 | 5 | 3 | 4 |
| Research Q2 | n=78 | n=47 | n=16 | n=15 | n=68 | n=46 | n=14 | n=15 | n=7 |
| Strongly agree | 32 | 18 | 9 | 5 | 21 | 18 | 0 | 6 | 1 |
| Agree | 14 | 9 | 3 | 2 | 10 | 17 | 2 | 1 | 3 |
| Neutral | 15 | 9 | 3 | 3 | 18 | 8 | 4 | 7 | 0 |
| Disagree | 9 | 5 | 1 | 3 | 10 | 0 | 3 | 0 | 1 |
| Strongly disagree | 8 | 6 | 0 | 2 | 9 | 3 | 5 | 1 | 2 |

**Table 2.** Factors associated with desire to learn of different incidental MRI findings. Values shown are odds ratio with 95% CI and p-values. Model for entirely benign findings not shown as it was not influenced by any factor other than racial/ethnic category.

|  | Past injury | Unclear findings | Serious findings with effective treatment | Serious findings with no effective treatment |
| --- | --- | --- | --- | --- |
| **Race** |  |  |  |  |
| South Asian | Reference | Reference | Reference | Reference |
| Hispanic | 0.99 (0.74, 1.32),<br>p=0.940 | 1.27 (0.96, 1.69),<br>p=0.096 | 1.16 (0.86-1.59),<br>p=0.318 | 0.99 (0.75-1.31),<br>p=0.947 |
| **White** | **1.24 (1.04-1.48),**<br>**p=0.017** | **1.44 (1.21, 1.73),**<br>**p<0.001** | **1.25 (1.04-1.51),**<br>**p=0.018** | **1.37 (1.15-1.62),**<br>**p<0.001** |
| Black | 1.05 (0.78-1.42),<br>p=0.751 | 1.07 (0.79, 1.44),<br>p=0.660 | 1.10 (0.80-1.51),<br>p=0.541 | 1.02 (0.76-1.37),<br>p=0.893 |
| **East Asian** | **0.84 (0.72-0.99),**<br>**p=0.037** | 0.93 (0.80, 1.10),<br>p=0.409 | 1.00 (0.85-1.17),<br>p=0.986 | 0.90 (0.77-1.05),<br>p=0.180 |
| **Age** |  |  |  |  |
| <50 | Reference | N.S. | Reference | Reference |
| 50-59 | 1.08 (0.88-1.34),<br>p=0.451 |  | 0.94 (0.77-1.16),<br>p=0.572 | 0.94 (0.76-1.16),<br>p=0.555 |
| **60-69** | 0.96 (0.81-1.14),<br>p=0.650 |  | 0.86 (0.73-1.02),<br>p=0.078 | **0.76 (0.64-0.89),**<br>**p=0.001** |
| **70+** | 0.83 (0.69-1.00),<br>p=0.052 |  | **0.77 (0.65-0.92),**<br>**p=0.005** | **0.70 (0.59-0.84),**<br>**p<0.001** |
| **Recruitment Site** |  |  |  |  |
| Events | N.S. | N.S. | Reference | N.S. |
| **Community Contacts** |  |  | **1.27 (1.09-1.49),**<br>**p=0.003** |  |
| Clinic |  |  | 1.14 (0.90-1.45),<br>p=0.261 |  |
| **Worry of future dementia/stroke** | **1.09 (1.02-1.16),**<br>**p=0.010** | **1.08 (1.01, 1.15),**<br>**p=0.025** | **1.08 (1.01-1.15),**<br>**p=0.024** | 1.06 (1.00-1.13),<br>p=0.058 |
| **MRI safe** | **1.09 (1.02-1.16),**<br>**p=0.012** | **1.07 (1.01, 1.14),**<br>**p=0.034** | N.S. | **1.10 (1.03-1.17),**<br>**p=0.003** |

There was no difference in perceived prevalence of incidental brain MRI findings (p=0.320), with most (76%) over-estimating the prevalence of incidental finding. A minority believed MRI could increase future cancer risks (12.5%; no difference by race/ethnicity, p=0.226). Compared to SA respondents, White respondents were more likely to have had any MRI (OR 2.91, 95% CI 1.29-6.58, adjusting for age and referral source; Table 1) or brain MRI (OR 3.82, 95% CI 2.20-6.62; Table 1).

### 3.2 Incidental findings on brain MRI

Most respondents (229/249, 92%) wished to learn about one or more incidental findings on their own brain MRI. SA and EA respondents expressed the lowest, while White respondents expressed the highest, desire to be informed of six incidental brain MRI findings (Fig 1). The greatest differences existed in benign as well as serious incidental findings. Relative to SA respondents, wishing to be informed of more incidental brain MRI findings was associated with White race (B=1.28, 95% CI 0.53-1.92, p<0.001) but not EA ethnicity. Age and recruitment site did not influence the desire to learn additional incidental findings.

**Fig 1.**
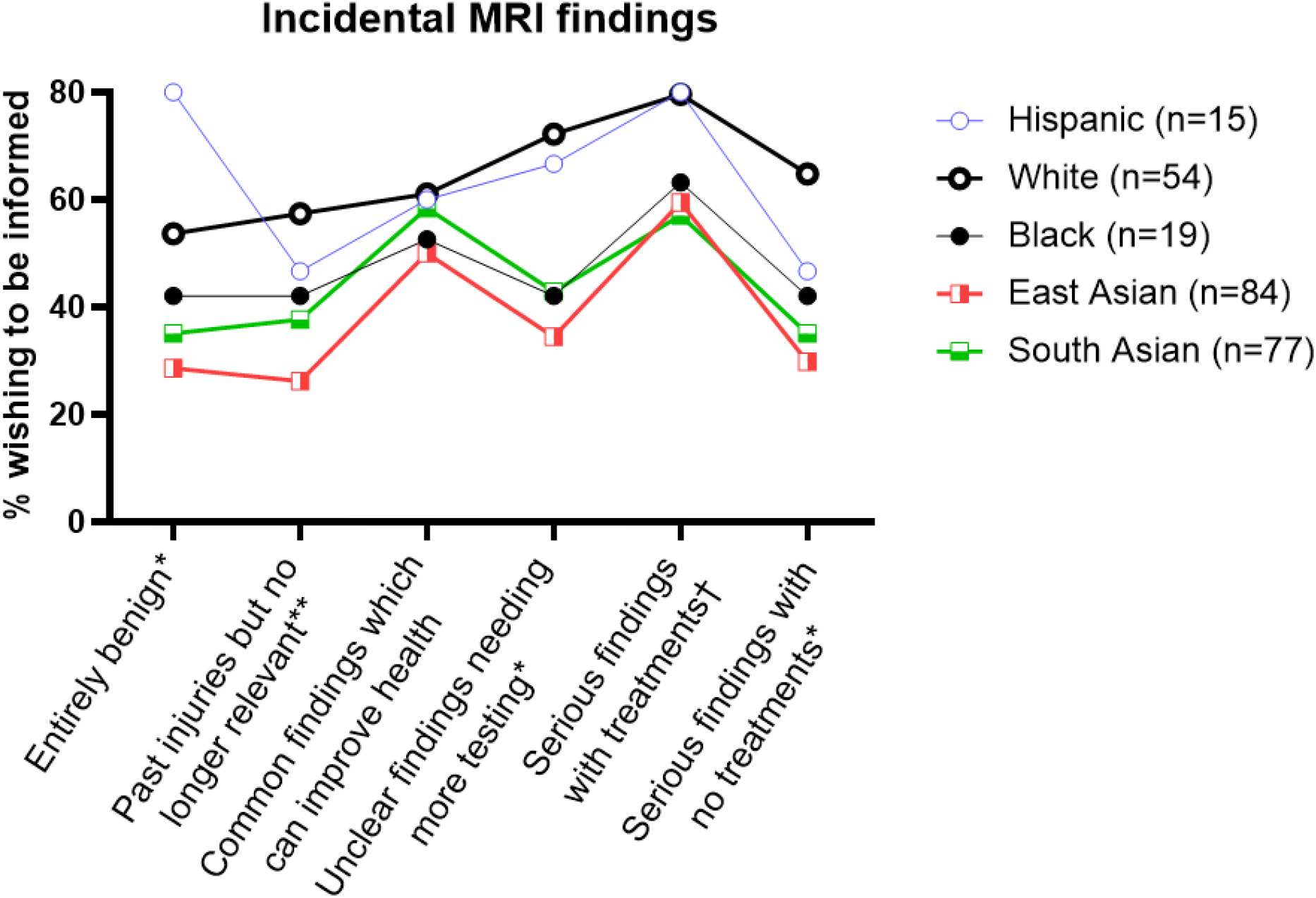
Proportion of respondents who wish to be informed of incidental brain MRI findings by types of finding (X-axis) and race/ethnicity. * p ≤ 0.001; ** p < 0.01, † p<0.05.

Of note, 35% of respondents did not wish to learn of serious MRI findings with effective treatments, and 52% did not wish to learn of uncertain findings requiring further testing.

### 3.3 Attitudes towards research participation, dementia, and MRI

208 participants (81%) provided complete responses to the ten Likert-scale questions. Concerning research participation, 139 (67%) disagreed with the statement *“Healthy people do not need to participate in medical research, because there is no direct benefit to them”* (Table 1, Research Q1), possibly reflecting the convenient nature of this cohort. Similarly, 115 (55%) agreed with the statement *“After my name is removed from my brain scan, I am comfortable with the idea of scientists (who are not all physicians) I have never met examine the images”* (Table 1, Research Q2). Compared to SA respondents, White respondents were more likely to believe healthy people should volunteer (P.O.=2.54, 95% CI 1.22-5.32, p=0.013) and share de-identified MRI for research (P.O.=2.33, 95% CI 1.45-3.74, p<0.001; adjusting for age). EA respondents also were more likely than SA respondents to believe healthy people should volunteer (P.O.=2.14 95% CI 1.13-4.05, p=0.019).

For the remaining Likert scale questions on attitudes related to brain health (Fig 2), PCA identified three PCs (Table 3): long-term MRI safety (PC1), internal LoC for developing dementia (PC2), and respondents’ own worries for future dementia/stroke risks (PC3). Compared to SA respondents, White respondents had lower internal LoC for dementia (p=0.043), and EA (p=0.006) and Hispanic (p=0.019) respondents had greater worries for future dementia/stroke risks. Respondents 50+ also believed in greater internal LoC than those <50 (p=0.026), but there was otherwise no difference among the older groups (50-59, 60-69, 70-79, 80+).

**Fig 2.**
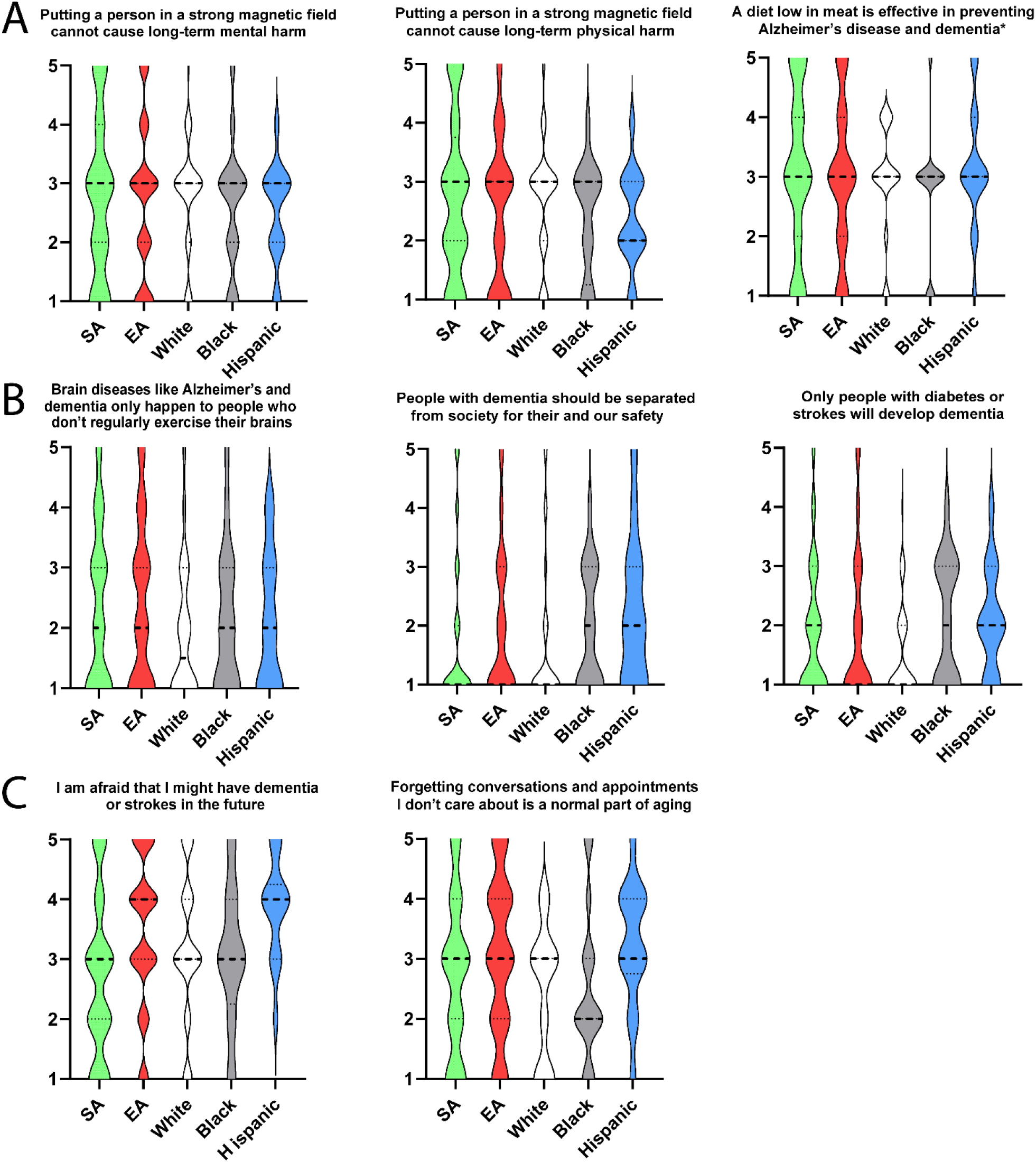
Attitudes related to brain health according to race/ethnicity and PCA. PC1 primarily involved long-term MRI safety (A, *low meat diet question had low loading overall but highest loading on PC1); PC2 involved internal LOC for developing dementia (B); PC3 involved worries for future dementia/stroke risks (C).

**Table 3.** Principal component analysis of attitudes related to brain health (loading scores > 0.100 shown). PC 1 related to long-term MRI safety, PC2 related to internal locus of control (LoC) for developing dementia, and PC3 related to respondents’ own worries for future dementia/stroke risks.

|  | PC1 | PC2 | PC3 |
| --- | --- | --- | --- |
| A strong magnetic field cannot cause long-term mental harm | 0.925 |  |  |
| A strong magnetic field cannot cause long-term physical harm | 0.921 | 0.103 |  |
| Diet low in meat prevents Alzheimer's and dementia | 0.293 | 0.185 | 0.235 |
| Brain diseases happen to people who don't exercise their brains |  | 0.829 | 0.113 |
| People with dementia should be separated from society |  | 0.743 |  |
| Only people with diabetes or strokes develop dementia |  | 0.664 |  |
| I am afraid that I might have future dementia or strokes |  |  | 0.834 |
| Forgetting things that are not important to me is normal aging | 0.102 | 0.149 | 0.720 |

### 3.4 Factors Associated with Favorable Research Attitudes

We next analyzed whether the three PCs might mediate the difference in research-related attitudes between SA and non-SA respondents.[29] Compared to SA respondents, we found White – but not EA – respondents to associate dementia risks with less internal LoC and this mediated the difference in attitudes towards healthy volunteers between the two groups (Fig 3A, 3B). In contrast, greater worry for future dementia/strokes was associated with more willingness to share de-identified MRI for research, but could not account for the greater willingness in White than SA respondents (Fig 3C, 3D).

**Fig 3.**
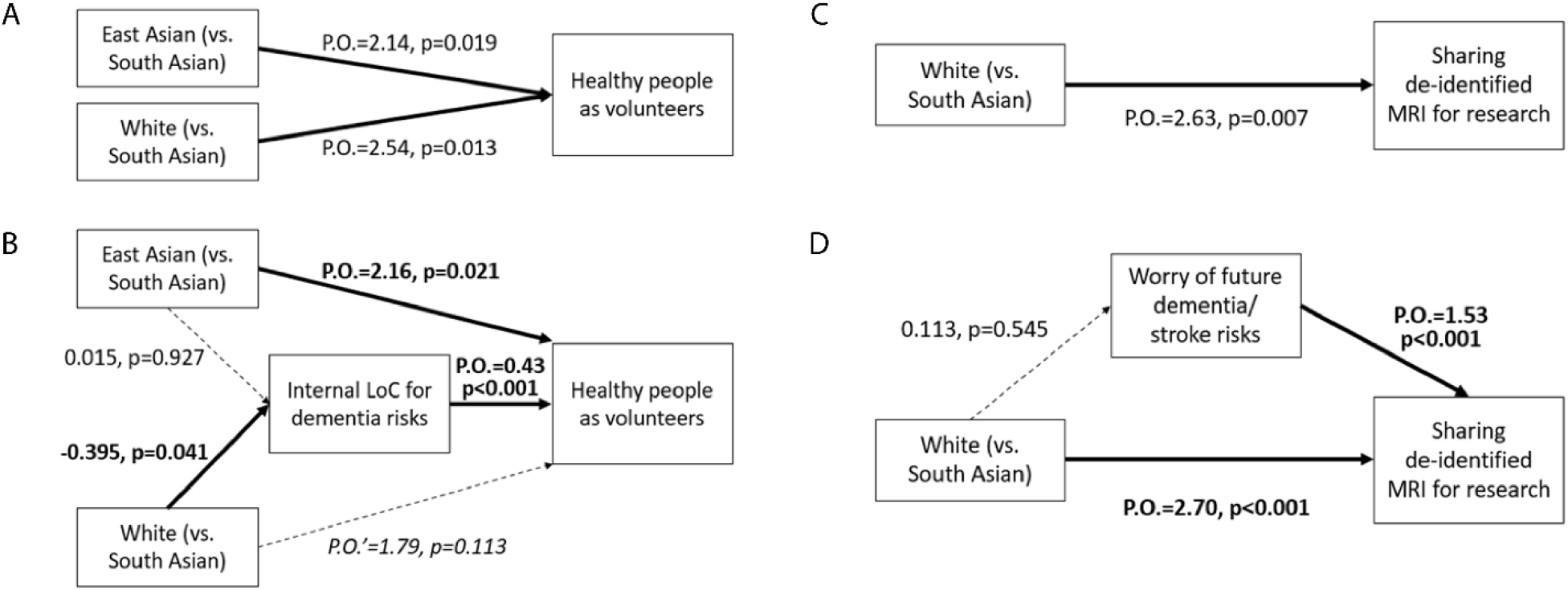
Mediation analysis for whether healthy people should volunteer in research (strongly disagree, disagree, neutral, agree, strongly agree) and sharing de-identified MRI for research (disagree, neutral, agree). Attitudes towards research participation were analyzed without (A, C) or with (B, D) the inclusion of attitudes related to brain health (internal LoC for dementia risks, fear for future dementia/stroke risks) as a mediator for difference between SA and other racial/ethnic respondents. P.O.: proportional odd.

### 3.5 Factors associated with disclosure of incidental MRI findings

Finally, we assessed whether MRI experience and attitudes related to dementia and research could explain differences in desire to learn of incidental MRI findings between racial/ethnic groups (Fig 1). Older age (60+) was associated with lower wish to learn of serious incidental findings regardless of treatment availability, and worry over future dementia/stroke risks was generally associated with greater desire to learn of uncertain as well as serious findings (Table 2). However, even after adjusting for age group, recruitment site, worry of future dementia/stroke, and perceived MRI safety, low desire among SA and EA respondents to learn of incidental MRI findings persisted (Table 2).

## 4. Discussion

Here we surveyed a cohort of older Asian and non-Asian Americans to better characterize their attitudes towards aging brain health research participation. We found both SA and EA respondents to have less interest in learning about incidental MRI findings than non-Asian respondents, and this difference was not influenced by prior MRI experience, perceived MRI safety, or dementia-related attitudes. SA respondents also showed relatively less support for healthy research volunteers and sharing their own de-identified MRI with researchers. Since disease- and experience-related factors poorly accounted for attitudes related to research between Asian and White adults, further studies are necessary to identify broader cultural and social factors which underlie these notable differences.

Aging studies involving EA[31] and SA [32] older adults are starting to generate epidemiological insight into psychosocial factors influencing brain health in these populations, but neuroimaging data – including willingness to undergo advanced brain imaging – are lacking.[33] With some exceptions,[34, 35] cross-racial/ethnic studies on dementia attitudes (not including those which only assessed one racial/ethnic group) have historically recruited mostly White, Black, and Hispanic participants. We did not find a difference in perception of MRI safety among racial/ethnic groups, but we found greater belief of internal LoC for dementia risks among SA respondents. We did identify greater worry for future dementia/stroke among EA respondents, in keeping with findings from small prior studies on EA immigrants and dementia caregivers.[36, 37] However, we found similarly low desire to learn about incidental MRI findings among EA and SA respondents. One potential explanation is that worries over future disease risks manifest differently between EA and SA adults. For example, whereas EA adults in diabetes prevention research noted wish to maintain social harmony as a barrier to participation,[38] SA adults would not participate due to a high belief in personal efficacy (e.g., can exercise diabetes away, so do not need to do prevention).[26] If so, “I don’t want to know” associated with fear and “I don’t need to know” associated with low self-perceived risks need to be better distinguished when considering disclosure of incidental findings in aging research.

It is further noteworthy that immigration history and the surrounding (“host”) culture can additionally influence older Asian adults’ behaviors. For example, one study found older Chinese adults’ dementia-related worries to correlate with knowledge among those living in Melbourne, but not those in Beijing.[39] Since past as well as current structural factors often regulate the regional entry of immigrant groups, studies involving EA and SA older adults in other U.S. regions will be necessary to pinpoint generalizable cultural and immigration-related elements underlying these group-level behavioral differences.

The variable desire to learn of incidental MRI findings among racial/ethnic groups underscores the urgency for thoughtful flexibility. Standards for disclosure of incidental findings in U.S. biomedical research abide by the ethical threshold of “actionability.”[40] Actionability can be determined according to well-established medical actions, patient/participant-initiated health-related actions, and life-plan decisions.[41] Challenges in a one-size-fit-all disclosure strategy in brain imaging were noted as early as 2002.[11, 18, 20] We could find no culturally-sensitive algorithm to determine if, how, and how many incidental findings from research MRI should be disclosed to older Asian adults living in the U.S. or U.K.; practices vary in Chinese aging studies without influence from prevalence of incidental findings.[42, 43] Multiple models have been proposed in disclosing genomic information to balance truth-telling and doing-no-harm.[44] An “ask-tell-ask” approach might be appropriate in MRI research involving healthy volunteers,[45] but stakeholder focus groups,[46] improved health literacy,[47] and incidental finding committees[48] are all potential solutions. Yet, none of these methods has been empirically tested in multi-cultural settings. It is also possible that, after trustworthy exposure to medical research and asymptomatic disease detection, those who choose not to learn of incidental findings will shift their preference in the same way the medical providers did for disclosing cancer diagnosis in the second half of the 20^th^ Century.[49] Therefore, picking a single convenient model may underestimate the cultural adaptability of Asian older adults and asymmetrically place the onus of informed consent on the research participants, even if a flexible or progressive disclosure model requires further evidence generation and refinement.

This study’s overall sample size is similar to previous ones analyzing disclosure of incidental MRI findings,[19, 50] but the convenience sample has a number of limitations. Importantly, we could not explain the different wishes for disclosure between Asian and White respondents despite assessing several attitude-related domains, nor why EA respondents had greater support for healthy people as research volunteers than SA respondents. We had to limit our surveys’ length based on Asian older adults’ hesitancy in research participation, at the cost of collecting more detailed demographic and socio-behavioral data. Gendered role (traditional and US-based), acculturation, socio-economic status (challenging to assess in both retired persons and immigrants), and prior employment in medicine or medical research are among factors which can influence attitudes. The geographic concentration of EA and SA adults in the NJ/NYC made this study feasible, but our findings may not generalize to other U.S. regions (especially those with different SA and EA immigration history). We performed PCA to derive summary measures related to attitudes, but we did not assess the Likert scale questions’ reliability over time. Finally, while Hispanic and Black respondents also showed potentially distinguishing trends, these two groups – as well as people of mixed or other race/ethnicity – were too small in number to explore potential causes for such trends.

In conclusion, we found similar preferences related to incidental MRI findings between SA and EA adults despite quite different support for research participation, internal LoC for dementia risks, and worries for future dementia/stroke. Based on these and prior findings, we caution researchers in generalizing the linkage between perceptions and behaviors from one ethnic group to another, and call for more flexible – and potentially individual-based – tailoring of incidental finding disclosure in MRI-related research.

## Data Availability

All data produced in the present study are available upon reasonable request to the authors

## Abbreviations

(EA): East Asian
(SA): South Asian
(LoC): Locus of Control
(PCA): Principal Component Analysis
(PC): Principal Component

## Acknowledgements

We thank the South Asian Total Health Initiative and RWJBarnabas Health Chinese Medical Program for their support in engaging older Asian adults.

